# Connecting proteomics and genomics to identify causal biomarkers specific for aortic valve stenosis

**DOI:** 10.1101/2025.11.27.25339923

**Authors:** Pardis Zamani, Ursula Houessou, Hasanga D. Manikpurage, Manel Dahmene, Zhonglin Li, Nathalie Gaudreault, Marie-Annick Clavel, Philippe Pibarot, Patrick Mathieu, Benoit J. Arsenault, Aida Eslami, Yohan Bossé, Sébastien Thériault

## Abstract

**Introduction:** Aortic valve stenosis (AS) is a progressive disease characterized by the calcification and narrowing of the aortic valve, leading to significant morbidity and mortality. Early detection and risk stratification remain a major clinical challenge. Identifying specific biomarkers could significantly improve risk prediction and disease management.

**Objective:** This study aims to (1) identify novel plasma protein biomarkers for incident AS, (2) establish causal relationships using genetic approaches, and (3) prioritize biomarkers specific to the aortic valve tissue.

**Methods:** We assessed the association between 2,923 unique plasma proteins measured using the Olink Explore assay and AS incidence in 52,632 UK Biobank participants with 487 incident AS cases over a median follow-up of 13 years. Multivariable Cox proportional hazards models were used to evaluate associations adjusted for cardiovascular risk factors. A stratified analysis was performed to investigate the association in men and women separately. For causal inference, we used protein quantitative trait loci (pQTL) Mendelian Randomization (MR). We then verified the aortic valve specificity of the proteins using our transcriptomic dataset of 500 human aortic valves. Using expression QTL (eQTL)-based MR, we evaluated the causal role of gene expression in AS and performed colocalization analyses.

**Results:** Our fully adjusted model identified 55 proteins significantly associated with AS incidence, with GDF15 showing the strongest association (HR=1.93 per SD, 95% CI: 1.67–2.23, P value = 3.3E-19). In sex-stratified analyses, 10 proteins showed a significant association with AS incidence in women, including 5 more strongly associated in women (CD38, CD80, IGFBPL1, NFASC, SERPINA9, P interaction <0.05), whereas 16 proteins were identified in men including 1 protein (REG1A) with a significantly stronger association in men. pQTL-MR suggested a potential causal role for 4 proteins (PCSK9, CHI3L1, NFASC, PRSS8). Transcriptomic integration confirmed high aortic valve expression for 11 candidates, with three genes demonstrating significant associations in aortic valve eQTL MR analyses, including LTBP2, for which the pQTL and eQTL colocalized.

**Conclusion:** The integration of proteomics and genomics allowed the identification of potential biomarkers and drug targets for AS, showing evidence of causality and tissue-specific expression.

## Introduction

Aortic valve stenosis (AS) is a prevalent and life-threatening cardiovascular condition. The pathogenesis of AS is multifactorial and involves lipid accumulation, oxidative stress, and inflammation^1^. These processes result in the fibrocalcific remodeling of aortic valve leaflets, ultimately leading to impaired blood flow^2,3^.

Currently, aortic valve replacement (AVR) is the only available treatment for AS, as no pharmacological agent has been proven effective in preventing or slowing disease progression. AVR was traditionally recommended for patients with severe symptomatic disease. Recent guidelines have expanded the indications for surgery in asymptomatic patients with severe AS^4–6^. In the absence of randomized clinical trials in symptomatic patients with moderate AS, there is an ongoing debate about the optimal management in this population^7^. Risk markers of disease progression could facilitate the selection of patients for intervention.

Only a few plasma biomarkers are currently recommended for the monitoring of AS in clinical practice, such as natriuretic peptides and high-sensitivity cardiac troponins, mostly to identify left ventricular dysfunction^4,5^. No biomarker specific to the AS disease process has sufficient supporting evidence to be used clinically. Circulating proteins involved in lipid metabolism, inflammation, tissue remodeling, hemostasis and mineralization have been associated with AS^8^. However, the value of these markers has not been thoroughly validated, and their specificity for the aortic valve has not been clearly established. Their relevance could also vary according to sex, considering previous evidence for the involvement of sex-specific processes in AS^9^.

Unbiased proteomic approaches provide an opportunity to identify novel biomarkers that may offer insights into disease mechanisms and serve as potential therapeutic targets. Integrating proteomic data with Mendelian Randomization (MR) can strengthen causal inference by leveraging genetic variants as instruments. Since genetic variants are randomly allocated at conception, MR minimizes biases from confounding and reverse causation, which are common in traditional observational studies^10^. This approach has been used successfully to identify genes with a potentially causal association between expression in aortic valve tissues and AS^11^.

The present study aims to identify plasma proteomic biomarkers associated with incident AS in a population-based cohort of 52,632 individuals and investigate sex-specific associations. By integrating proteomic profiling with MR analysis and aortic valve transcriptomics, we seek to identify specific circulating proteins that are causally linked to AS and may serve as potential biomarkers or therapeutic targets.

## Methods

### UK Biobank

Between 2006 and 2010, the UK Biobank (UKB) recruited over 500,000 participants aged 40–69 years from across the United Kingdom^12^. UKB received approval from the British National Health Service, Northwest - Haydock Research Ethics Committee (16/NW/0274). All participants provided informed consent. Participants underwent extensive health assessments, including comprehensive questionnaires, physical measurements, blood sample collection, and long-term health follow-up through linkage to electronic health records. The UKB data are integrated with national registries, including hospital patient records, cancer and death registries, and primary care records. This research has been conducted using the UK Biobank Resource under application number 25205.

### Proteomic data

We used proteomic data from the UK Biobank Pharma Proteomics Project (UKB-PPP), which includes a subset of UKB participants (n = 53,014). Using the antibody-based Olink Explore 3,072 Proximity Extension Assay technology, 2,923 unique proteins were measured in plasma across eight protein panels (cardiometabolic, cardiometabolic II, inflammation, inflammation II, neurology, neurology II, oncology, and oncology II). Protein concentrations were log2 transformed and normalized. Details on measurement, quality control procedures, and validation are described elsewhere^13^.

### Incident AS

The study outcome, incident AS, was defined as a first-time diagnosis recorded after recruitment based on the International Classification of Diseases (ICD) codes I35.0 or I35.2. Individuals with a AS diagnosis prior to recruitment or a history of rheumatic heart disease (I00–I02, I05–I09) were excluded. Individuals who underwent replacement or repair of the aortic valve (OPCS-4 codes K26/K30.2) or with self-reported AS were excluded if they did not have a diagnosis of incident AS (Supplementary Figure 1).

### Cox regression hazard ratio model

We constructed multivariable Cox proportional-hazards models to investigate the association between plasma proteins and the incidence of AS. Participants were censored at the time of loss to follow-up, death or at the end of the follow-up period (31 October 2022) if they were still alive and did not develop AS. Participants diagnosed with AS within 3 months after baseline were excluded from the analysis.

Two models were constructed for analysis. The base model was adjusted for age and sex. Subsequently, a fully adjusted model was developed, incorporating additional cardiovascular risk factors, including body mass index (BMI), smoking status, estimated glomerular filtration rate (eGFR), genetic ancestry (Caucasian or not), systolic blood pressure (SBP), and low-density lipoprotein cholesterol (LDL-C). SBP values were adjusted by adding 15 mmHg for individuals on anti-hypertensive medication, and LDL-C levels were corrected by dividing by 0.7 for individuals on lipid-lowering therapy, to account for the influence of treatment^14,15^. eGFR was calculated using the Chronic Kidney Disease Epidemiology Collaboration (CKD-EPI) equation.

Information on smoking and ethnicity was collected through patient self-reports. Smoking status was classified into three categories: current smoker, former smoker, and never smoker. Ethnicity was categorized into five groups from the response to the questionnaire: White, Black, Asian, Mixed and Other ethnic group. BMI was calculated using height and weight measured at the baseline visit.

Among the included 52,632 UKB-PPP participants, missing data were observed at 255 (0.48%) for BMI, 2,829 (5.4%) for SBP, 2,575 (4.9%) for eGFR, and 2,665 (5.1%) for LDL-C. To address this, we imputed the missing values using linear regression models, with age, sex, ethnicity, and the first 10 principal components of genetic ancestry as predictors. We then performed analyses stratified by sex adjusting for the covariates in the second model. Correction for multiple testing was performed using a Bonferroni adjustment; proteins with a P value < 1.7E-5 were selected for further analysis. The proportional hazards assumption for the Cox regression analysis was visually inspected by using Schoenfeld residuals plots. To additionally evaluate potential sex-specific differences in the associations, interaction terms for plasma protein levels and sex were included in the fully adjusted model. This allowed us to assess whether the relationship between plasma proteins and AS differed significantly between women and men.

### Enrichment analysis

To elucidate the biological mechanisms underlying the proteins identified as associated with AS incidence, pathway enrichment analyses were conducted using Metascape^16^. A custom background of 2,923 genes corresponding to the proteins in UKB-PPP was defined. To explore sex-specific biological differences, the pathway analysis was performed separately for proteins showing differential associations (P value < 0.05 for interaction) between men and women, grouped by sex based on effect size. Default settings were applied, including a minimum overlap of three genes per pathway, and a p-value cutoff of 0.01.

### Proteome-wide Mendelian randomization

We performed MR analyses to investigate whether the identified proteins in Cox regression models may have a causal role in the incidence of AS. To avoid potential bias, two distinct genetic summary statistics datasets of European ancestry were used in this analysis for exposure and outcome. The primary data for genetic instruments for plasma protein levels was extracted from the deCODE dataset. The deCODE genome-wide association study (GWAS) measured 4,907 aptamers in 35,559 Icelanders using the SomaScan assay (SomaLogic). The signal was normalized by rank-inverse normal transformation^17^. Genetic variants located within 500 kilobases (kb) of the transcriptional start and end sites of the gene encoding the corresponding protein were selected as instruments based on their association with protein level at genome-wide significance (P value < 5E-8). Summary statistics for AS were obtained from a GWAS meta-analysis comprising 14,819 cases and 927,044 controls of European ancestry, as described by Thériault et al. (2024)^11^.

Proteins encoded by genes located on the X and Y chromosomes were excluded, as genetic data for these chromosomes were not available in the GWAS mentioned above. To ensure independence, linkage disequilibrium (LD) clumping was performed using an r² threshold of 0.1 within a 1 Mb window using the clump function implemented in PLINK v1.9^18^, with LD information obtained from individuals of European ancestry from the 1000 Genomes Project. A minimum of 3 independent cis-genetic variants was required. The primary analysis was conducted using the inverse variance-weighted (IVW) method with a significance threshold of P value < 0.05. Additional sensitivity analyses including weighted median estimation^19^ (P value < 0.05 for robust effects) and MR-Egger regression^20^ (P intercept ≥ 0.05) were carried out.

### RNA-seq

To assess the aortic valve specificity of plasma proteins associated with AS incidence, we verified their corresponding gene mRNA levels in our local transcriptomic data from human aortic valve tissues^11^. Human aortic valve tissue samples were obtained from participants recruited at the Institut universitaire de cardiologie et de pneumologie de Québec-Université Laval (IUCPQ-ULaval) between 1998 and 2019. The data includes 500 samples of healthy and diseased human aortic valves from individuals of European ancestry. Healthy aortic valves (n = 60) were obtained from heart transplant recipients, while diseased aortic valves (n = 440) were collected from patients undergoing aortic valve replacement (AVR). The study was approved by the Ethics Committee of the IUCPQ–ULaval, and all participants provided written informed consent prior to enrollment.

Echocardiographic assessments were performed to evaluate the hemodynamic severity of aortic valve stenosis, classifying cases as very severe, severe, moderate, or mild/none^4^. Bulk RNA-sequencing was performed on an Illumina NovaSeq 6000 instrument aiming for >50 million paired-end reads per sample^11^ and quality control was done by FastQC v0.11.5 and MultiQC v1.10 applications^21^. Reads were collapsed into a single transcript model using GENCODE Release 41 (GRCh38) as the reference. RNA-SeQC 2^22^ was used to obtain read counts and Transcripts Per Million (TPM) values. We defined genes with high expression in the aortic valve if the expression was above the 90th percentile of all genes, corresponding to >78 TPM^11^.

### Aortic valve transcriptome-wide Mendelian Randomization

To investigate whether genetically predicted aortic valve gene expression influences AS risk, we performed expression quantitative trait loci (eQTL)-based Mendelian randomization for the proteins with significant association in the fully adjusted Cox model and high gene expression in human aortic valve tissues. To identify *cis*-expression quantitative trait loci (*cis*-eQTLs), we analyzed genetic variants within a 1 Megabase (Mb) window of the genes on chromosomes 1 through 22. The analysis was adjusted for covariates, including age, sex, current smoking status, the top 60 Probabilistic Estimation of Expression Residuals (PEER) factors, and the first five genetic principal components. Variants with a minor allele frequency (MAF) ≥ 0.01 and an imputation quality score ≥ 0.3 were included. QTLtools v1.1 was used to assess the associations between genetic variants and gene expression. Statistically significant eQTLs were identified using a false discovery rate (FDR) P value threshold of < 5%^11^. For each eligible gene, we extracted cis-eQTLs within ±500 kb of the transcription start site. To ensure instrument independence, clumping was performed using an r² threshold of < 0.1. Summary statistics for SNP–AS associations were obtained from the GWAS meta-analysis described above^11^. Our primary analysis used IVW regression. To evaluate the presence of horizontal pleiotropy and the robustness of the findings, sensitivity analyses were performed using MR-Egger regression (P value intercept ≥ 0.05) and the weighted median method (P value < 0.05). All MR analyses were conducted in R version 4.4.1 using the MendelianRandomization v.0.9.0 package.

### Colocalization analysis

To determine whether the variants associated with gene expression and those associated with circulating protein levels were the same, we performed Bayesian colocalization analyses integrating aortic valve tissue eQTLs with pQTLs from the UKB-PPP^13^ and aorta tissue eQTLs from GTEx (v8)^23^. As some corresponding pQTLs were not available in the deCODE dataset, the UKB-PPP was selected for this analysis; furthermore, it utilizes the same Olink platform for protein measurement, ensuring methodological consistency. For each protein-coding gene that showed significant associations in our eQTL-MR analysis (P value < 0.05), we examined a window of ±500 kb on each side of the gene. Using COLOC v3.2.1, we calculated posterior probabilities for four competing hypotheses: shared causal variants (PP.H4), distinct causal variants (PP.H3), single association only (PP.H1/H2), and no association (PP.H0). PP.H4 > 0.75 was considered strong evidence for colocalization.

### Protein Druggability

To evaluate the therapeutic potential of our identified protein biomarkers, we performed a druggability analysis using the Drug-Gene Interaction Database (DGIdb, v5.0, www.dgidb.org). This resource integrates drug-gene interactions, text-mining, and clinical relevance from multiple curated sources, including DrugBank, PharmGKB, ChEMBL, and Drug Target Commons. We prioritized drugs with an interaction score ≥ 0.1.

## Results

### Baseline characteristics of the study participants

Our study population included 52,632 UKB-PPP participants. Among them, 54.1% were female and 45.9% were male. Baseline characteristics of the overall study population stratified by AS incidence are presented in Table 1. Baseline characteristics stratified by sex are provided in Supplementary Table 1. Overall, women exhibited a more favorable cardiovascular risk factor profile compared to men, characterized by lower BMI and SBP, as well as a lower prevalence of diabetes, smoking, hypertension, and coronary artery disease (CAD).

**Table 1:** Baseline characteristics of the study population. *Corrected for antihypertensive medication (+15 mmHg) ‡Corrected for lipid-lowering therapy (/0.7)

| Characteristics | Without AS<br>(n=52,145) | With incident AS<br>(n= 487) |
| --- | --- | --- |
| Age, years, mean (SD) | 56.7 (8.2) | 63.1 (5.3) |
| Sex, Women, n (%) | 28271 (54.2) | 180 (36.9) |
| BMI, kg/m<sup>2</sup>, mean (SD) | 27.4 (4.8) | 30.1 (5.5) |
| Smoking, n (%) |  |  |
| Current | 5,498 (10.5) | 65 (13.3) |
| Former | 18,077 (34.7) | 230 (47.2) |
| Blood pressure, mmHg, mean (SD) |  |  |
| Systolic blood pressure* | 140.9 (20.8) | 154.3 (21.6) |
| Diastolic blood pressure | 82.1 (10.1) | 82.1 (11.2) |
| Blood lipids, mmol/L, mean (SD) |  |  |
| LDL cholesterol‡ | 3.7 (0.8) | 3.9 (0.9) |
| HDL cholesterol | 1.4 (0.4) | 1.3 (0.4) |
| Total cholesterol | 5.6 (1.1) | 5.4 (1.3) |
| Triglycerides | 1.7 (1.0) | 2.0 (1.2) |
| eGFR, mL/min/1.73 m<sup>2</sup>, mean (SD) | 90.1 (14.2) | 82.2 (17.4) |
| Diabetes, n (%) | 5,056 (9.7) | 141 (28.9) |
| CAD, n (%) | 6,130 (11.7) | 273 (56.1) |
| Hypertension, n (%) | 17,482 (33.5) | 382 (78.4) |
| Ethnicity, n (%) |  |  |
| White | 48,599 (93.2) | 471 (96.7) |
| Asian | 1,117 (2.1) | 10 (2.0) |
| Black | 1,211 (2.3) | 3 (0.6) |
| Mixed | 345 (0.6) | 1 (0.2) |
| Others | 615 (1.2) | 2 (0.4) |

### Cox regression

Over a median follow-up of 13 years, 487 participants developed AS (36.9% women). We evaluated the association between plasma levels of 2,923 distinct proteins and the risk of AS using Cox proportional hazards models. After accounting for multiple testing, 424 proteins were associated with the risk of AS in the base model adjusted for age and sex (P value < 1.7E-5), of which 406 were positively associated and 18 were negatively associated with incident AS (Figure 1a and Supplementary Table 2). Growth differentiation factor 15 (GDF15) showed the strongest association with a hazard ratio (HR) of 2.25 per standard deviation (SD) increase in protein level (95% CI: 2.02–2.52, P value = 3.79E-46). In the fully adjusted model, which accounted for age, sex, BMI, smoking, eGFR, genetic ancestry, SBP, and LDL-C, 55 proteins demonstrated a significant association with incident AS. GDF15 remained the most strongly associated with AS (HR = 1.93 per SD, 95% CI: 1.67–2.23, P value = 3.26E-19) (Figure 1b<u>, and</u> Supplementary Table 3).

**Figure 1:**
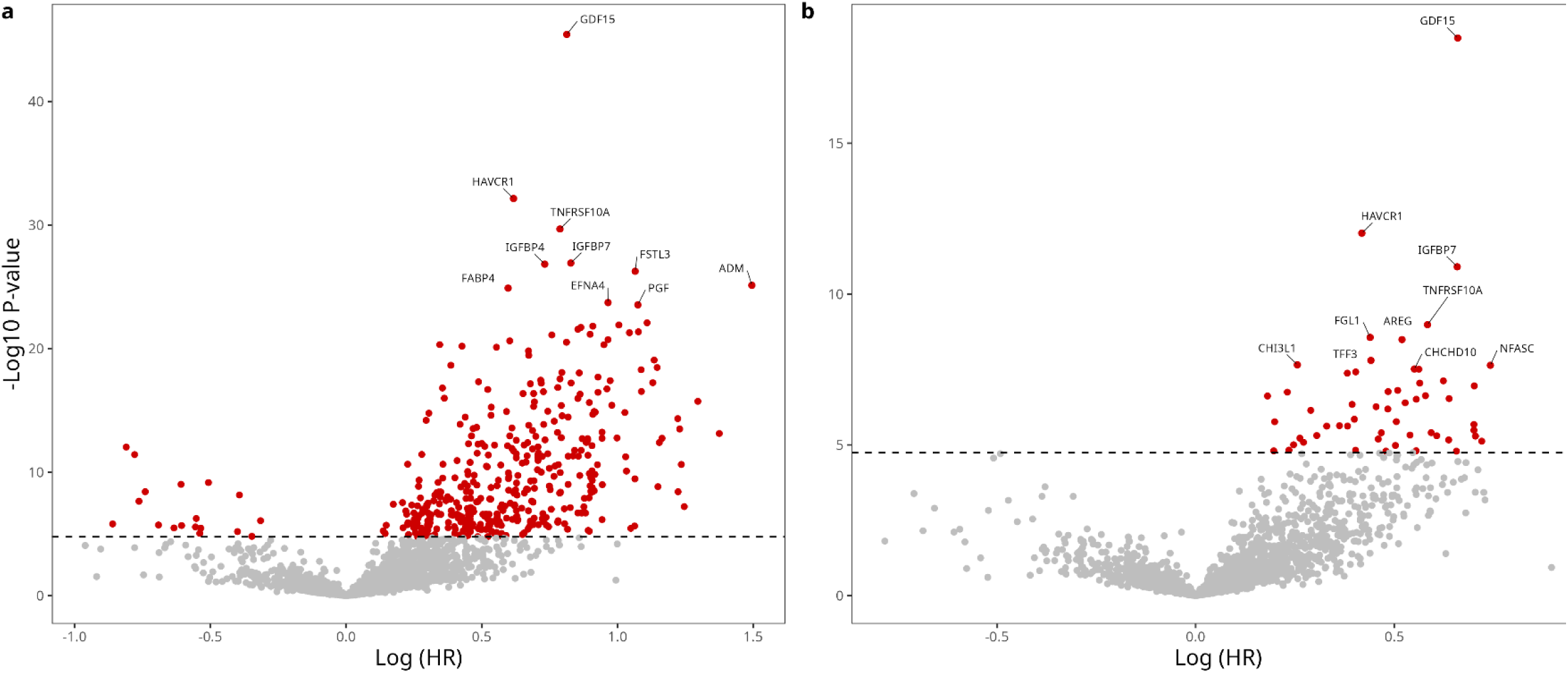
Volcano plots representing the association between plasma proteins and incident aortic valve stenosis (AS). The x-axis shows the log-hazard ratio (HR) from Cox proportional hazards regression per standard deviation increases in protein levels, and the y-axis displays –log P-values. Panel (a) shows results from the base model, adjusted for age and sex (P value threshold = 1.71E-5), while panel (b) shows the fully adjusted model, including age, sex, body mass index, smoking status, estimated glomerular filtration rate, genetic ancestry, systolic blood pressure, and low-density lipoprotein cholesterol (P value threshold = 1.71E-5). Horizontal dashed lines indicate the significance threshold. Proteins reaching significance (Bonferroni corrected P < 0.05) are highlighted (red), with selected top 10 hits labeled; gray points indicate non-significant associations.

To explore the biological pathways linking the identified proteins in the adjusted model, we conducted enrichment analysis, which revealed inflammatory signaling (TNFs bind their physiological receptors) as the most statistically significant term. Other enriched pathways included activation of NF-kappaB-inducing kinase activity, cell surface receptor protein tyrosine kinase signaling pathway, and intrinsic apoptotic signaling pathway (Supplementary Table 4).

Stratified analyses revealed notable sex-specific associations. In women (n = 28,451), 10 proteins reached statistical significance, with IGFBP7 displaying the most significant effect (HR = 2.25 per SD, 95% CI: 1.70–2.98, P value = 1.49E-08) (Figure 2a). In men (n = 24,181), 16 proteins were significantly associated with AS incidence, with GDF15 showing the strongest association (HR = 2.06 per SD, 95% CI: 1.73–2.45, P value = 8.09E-16) (Figure 2b).

**Figure 2:**
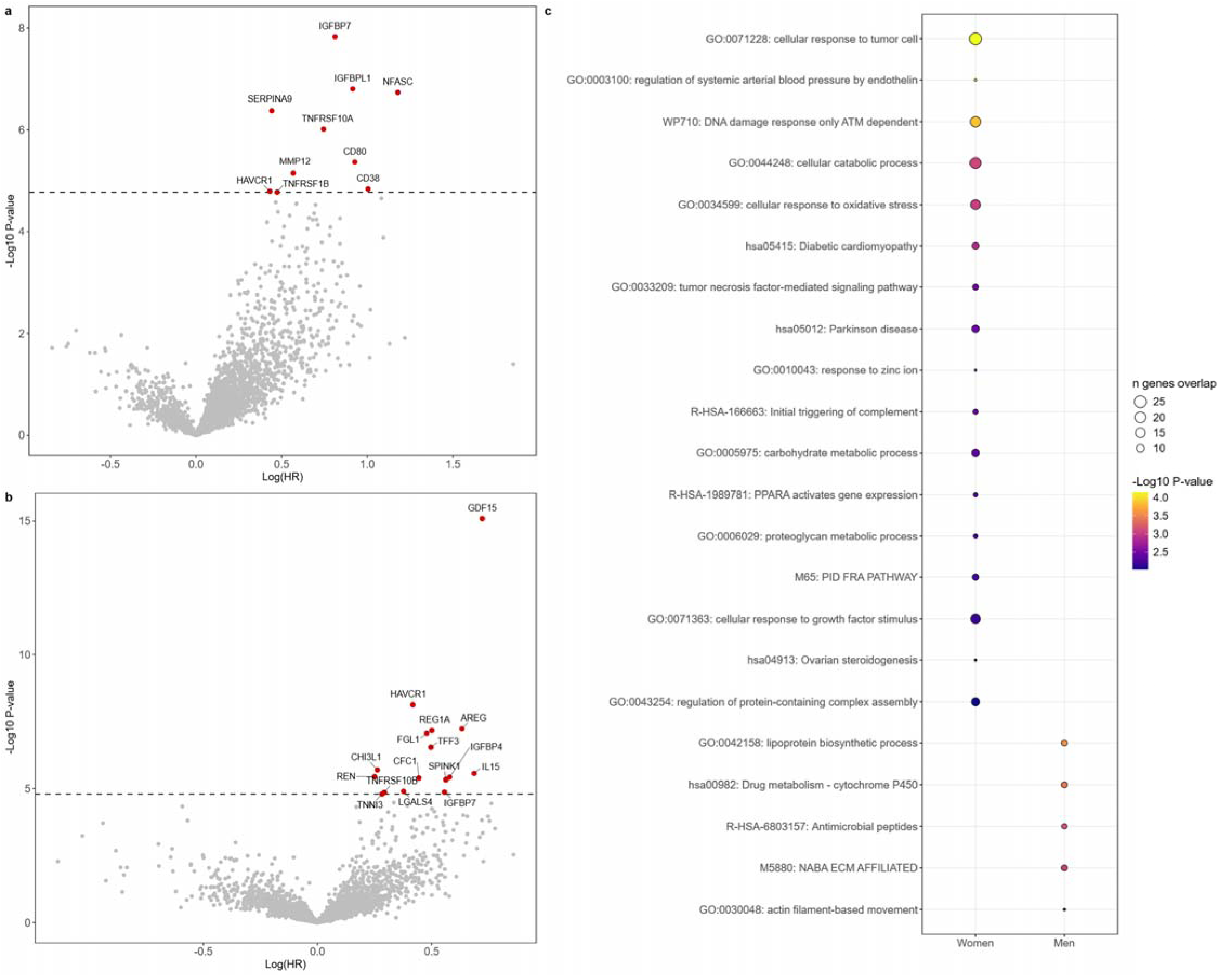
Plasma proteins associated with incident AS in sex-stratified analyses. Volcano plot of significant protein-AS associations from Cox proportional hazards regression in (a) women and (b) men. Proteins passing the significance threshold (P value < 1.71E-5) are labeled (in red). Hazard ratios (HR) represent risk per standard deviation increase in protein level. (c) Bubble plot of enriched pathways from sex-stratified analyses. Bubble size represents the number of proteins per pathway; color indicates the −log (P value) for pathway enrichment. Pathways were derived from Metascape analysis using proteins with sex-specific interactions (P interaction < 0.05).

Among the significant proteins, IGFBPL1, NFASC, SERPINA9, TNFRSF10A, CD80, MMP12, CD38, and TNFRSF1B were exclusively found in women, whereas GDF15, AREG, REG1A, FGL1, TFF3, CHI3L1, IL15, REN, IGFBP4, CFC1, SPINK1, LGALS4, TNFRSF10B, and TNNI3 were uniquely associated with AS in men, highlighting potential sex-specific biological mechanisms in disease development (Supplementary Tables 5 and 6). Interaction analyses confirmed significant sex differences for 172 proteins, suggesting differential biological roles of these circulating factors between women and men (Supplementary Table 7). To perform separate pathway enrichment analyses, proteins with significant interaction terms were stratified based on the magnitude of their effect estimates in women and men, respectively. Among women, the top enriched pathways included regulation of systemic arterial blood pressure, cellular response to oxidative stress, and diabetic cardiomyopathy. In men, the proteins exhibited significant enrichment in lipoprotein biosynthetic process, NABA ECM AFFILIATED, and actin filament-based movement (Figure 2c). We integrated findings from sex-stratified Cox proportional hazards models with results from sex interaction analyses (P interaction < 0.05). This approach identified 6 circulating proteins that met both criteria: (1) significant associations with incident AS in either men or women after full covariate adjustment, and (2) statistically significant differences in effect between sexes. These 6 proteins included 1 male-specific (REG1A) and 5 female-specific associations (CD38, CD80, IGFBPL1, NFASC, SERPINA9) (Figure 3).

**Figure 3:**
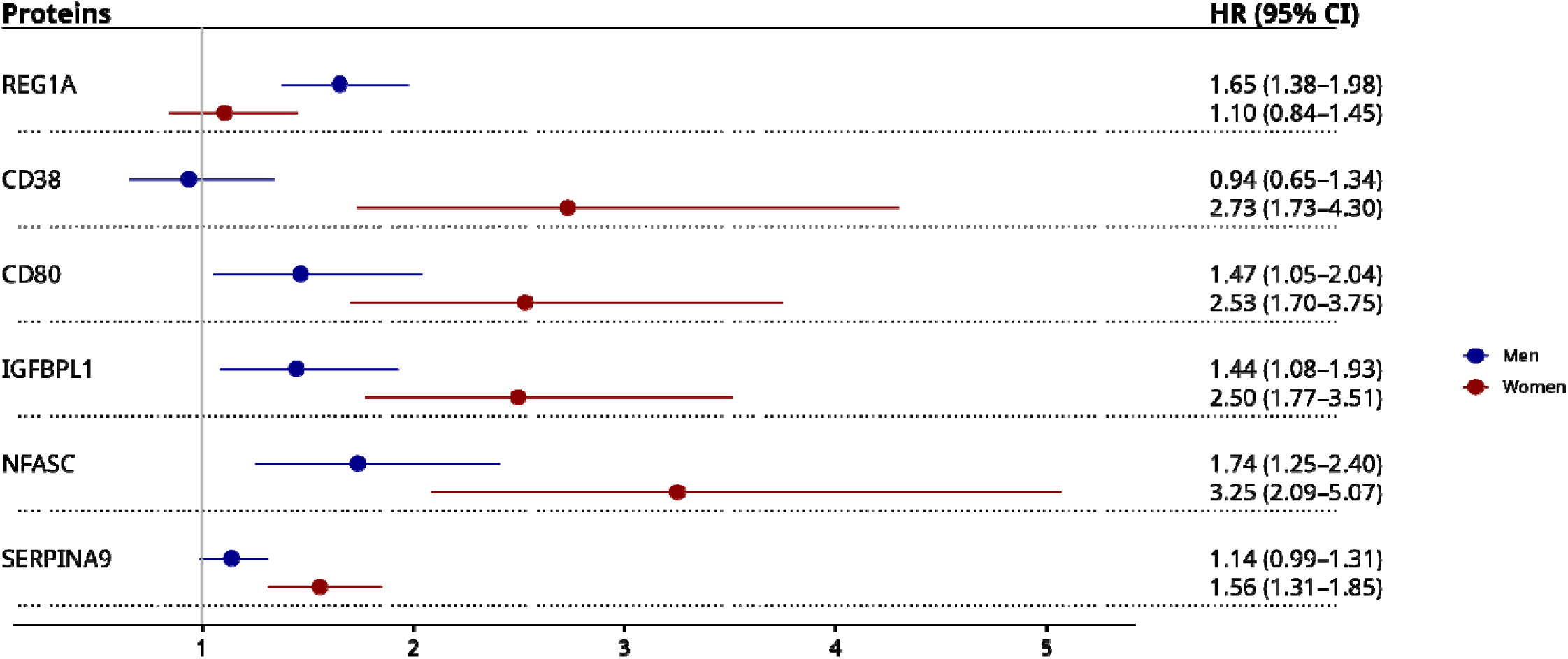
Plasma proteins associate with incident AS with significant sex interaction. The forest plot displays hazard ratios (HR) per SD and 95% confidence intervals for six proteins showing both significant associations in sex-stratified Cox proportional hazards regression (fully adjusted model), and significant sex interaction (P < 0.05). Results for men are shown in blue, results for women are shown in red. REG1A demonstrated men-specific association, while CD38, CD80, IGFBPL1, NFASC, and SERPINA9 showed women-predominant associations. The vertical line indicates null effect (HR = 1).

### pQTL MR analysis

We performed Mendelian randomization analyses to assess the causal effects of circulating proteins on AS risk. Among the 424 proteins identified as significant in our base model, 325 were available in the deCODE dataset. After clumping (LD r² < 0.1 within a 1 Mb window), 180 proteins with at least 3 genetic variants selected were included in the analysis. Using IVW as the primary method, MR identified 43 proteins with nominal evidence of a potential causal association (P < 0.05). Sensitivity analyses indicated that 38 proteins showed no strong evidence of horizontal pleiotropy based on the MR-Egger intercept test (P ≥ 0.05). Among these, 16 proteins also showed significant association using the weighted median method (P < 0.05), supporting the robustness of the observed effects (Supplementary Table 8). Among the 55 proteins in the fully adjusted model, 4 proteins exhibited robust and significant association in MR (Figure 4). The strongest association was observed for Proprotein Convertase Subtilisin/Kexin type 9 (PCSK9), where increase in genetically predicted levels was associated with higher AS risk (OR = 1.18 per SD, 95% CI: 1.12–1.24, P value = 2.66E-10). Additional positive associations were observed for chitinase-3-like protein 1 (CHI3L1; OR = 1.03 per SD, 95% CI: 1.02–1.05, P value = 6.42E-5), neurofascin (NFASC; OR = 1.06 per SD, 95% CI: 1.02–1.10, P value = 0.005), and prostasin (PRSS8; OR = 1.32 per SD, 95% CI: 1.07–1.64, P value = 0.01) (Supplementary Table 8).

**Figure 4:**
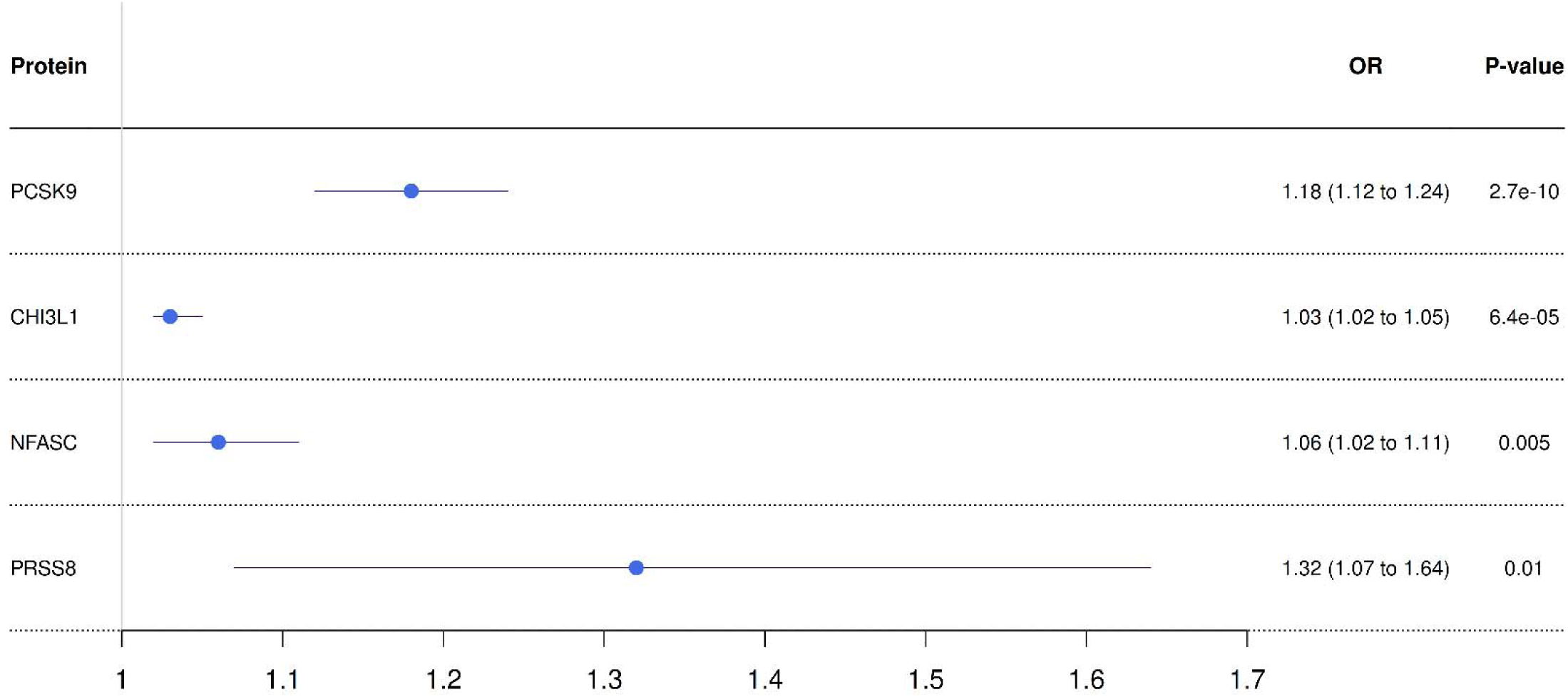
Plasma proteins associated with AS in Mendelian randomization analyses. The forest plot shows the causal effect estimates from pQTL Mendelian randomization analyses. Four proteins show significant associations with AS risk among the proteins significant in the Cox proportional hazards regression fully adjusted model. Error bars represent 95% confidence intervals for the inverse-variance weighted (IVW) estimates.

### RNA expression

To evaluate the aortic valve specificity of significant plasma proteins associated with AS incidence in our fully adjusted model, we examined their corresponding gene expression levels in human aortic valves. Out of these 55 proteins, several exhibited significantly elevated mRNA levels in aortic valve tissue (defined as >90th percentile of all genes corresponding to >78 TPM): IGFBP7, B2M, C7, IGFBP4, LTBP2, COL4A1, GGT5, TNFRSF11B, SHISA5, CHCHD10, and ITGBL1 (Supplementary Table 9).

### eQTL MR

Using cis-eQTLs located within ±500 kb of the transcription start site, we performed Mendelian Randomization for the 11 selected highly expressed genes. Among the genes tested, *COL4A1, LTBP2* and *B2M* showed statistically significant genetic associations with AS risk. Higher genetically predicted expression of *COL4A1* (OR = 1.32 per SD, 95% CI: 1.10 – 1.59, P value = 0.003), *LTBP2* (OR = 1.21 per SD, 95% CI: 1.05 – 1.40, P value = 0.01), and *B2M* (OR = 1.21 per SD, 95% CI: 1.07 – 1.36, P value = 0.02) were each robustly associated with an increased risk of AS with no evidence of pleiotropy (Figure 5).

**Figure 5:**
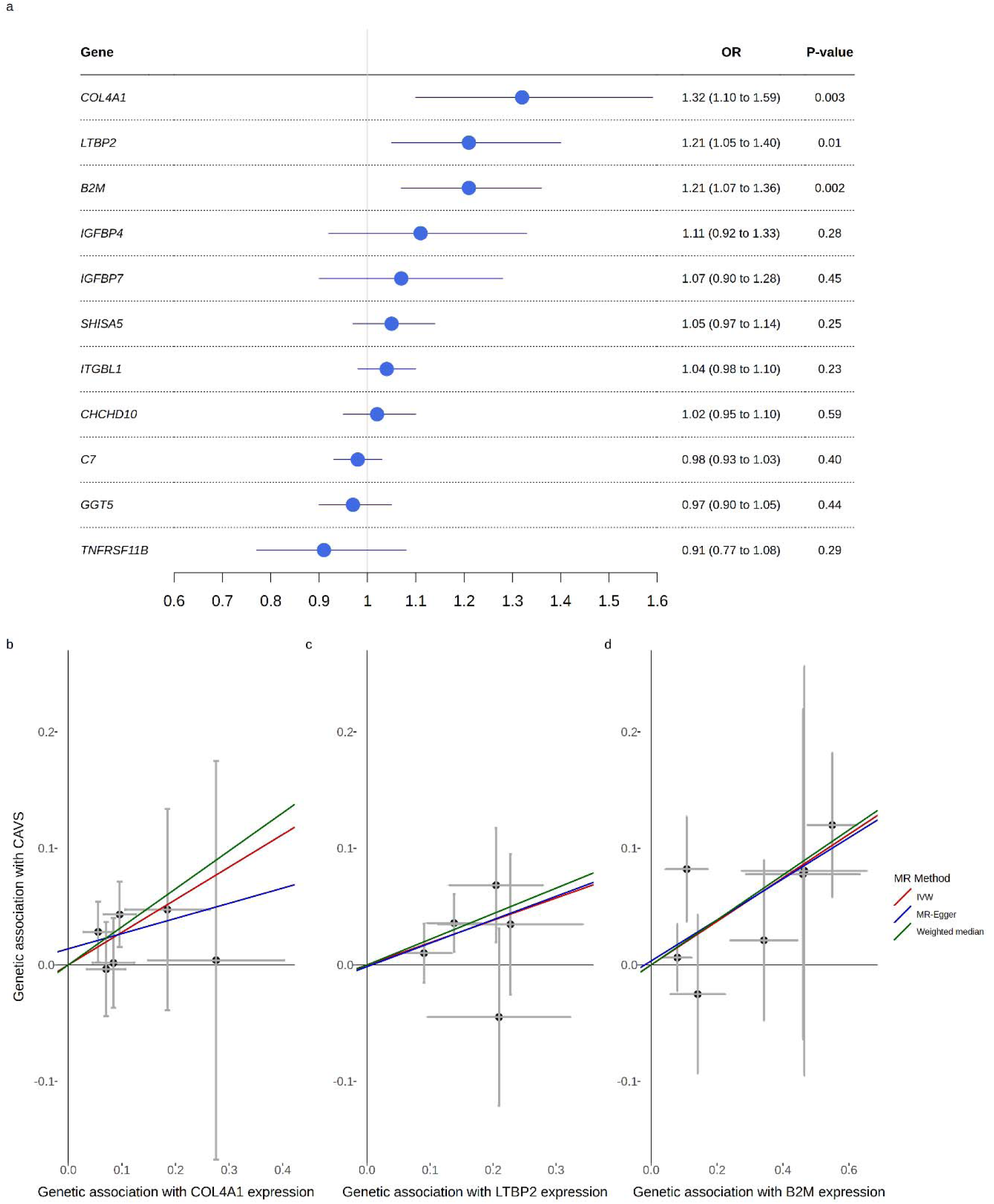
Mendelian randomization analyses in aortic valve tissues. (a) Forest plot depicting the causal effect estimates from eQTL Mendelian randomization analyses, including the 11 proteins with high expression levels in human aortic valve tissues. Causal estimates were derived using the inverse-variance weighted (IVW) method with valve tissue cis-eQTLs (±500 kb). *COL4A1*, *LTBP2* and *B2M* demonstrated statistically significant associations with AS risk. Error bars represent 95% confidence intervals for the IVW estimates. (b, c, d) Mendelian randomization scatter plots showing the association between genetic variants influencing (b) *COL4A1*, (c) *LTBP2*, and (d) *B2M* expression in aortic valve tissues (x-axis, eQTL effect) and their corresponding effect on AS risk (y-axis, GWAS effect). Each point represents an independent cis-eQTL (r² < 0.1). Colored lines indicate Mendelian randomization estimates: Inverse-variance weighted (red), Egger (blue), and weighted median (green).

### Colocalization analysis

For *COL4A1*, *LTBP2* and *B2M*, the three genes showing significant associations in our eQTL-based MR, we performed colocalization analyses (Supplementary Table 10). For *LTBP2*, we found strong evidence of colocalization between aortic valve eQTLs and plasma pQTLs (PP.H4 = 0.93), indicating that the same variants influence both *LTBP2* expression in valve tissue and its circulating protein levels. We also found that the lead pQTL is a significant eQTL in the aorta from GTEx and that the signals colocalized between eQTL in aortic valve and eQTL in aorta (PP.H4 = 0.88) (Figure 6). In contrast, *COL4A1* and *B2M* showed no evidence of colocalization between pQTLs and aortic valve eQTLs (PP.H4 < 0.5) (Supplementary Figures 2 and 3). No significant eQTLs were found in aortic tissues from GTEx for these genes.

**Figure 6:**
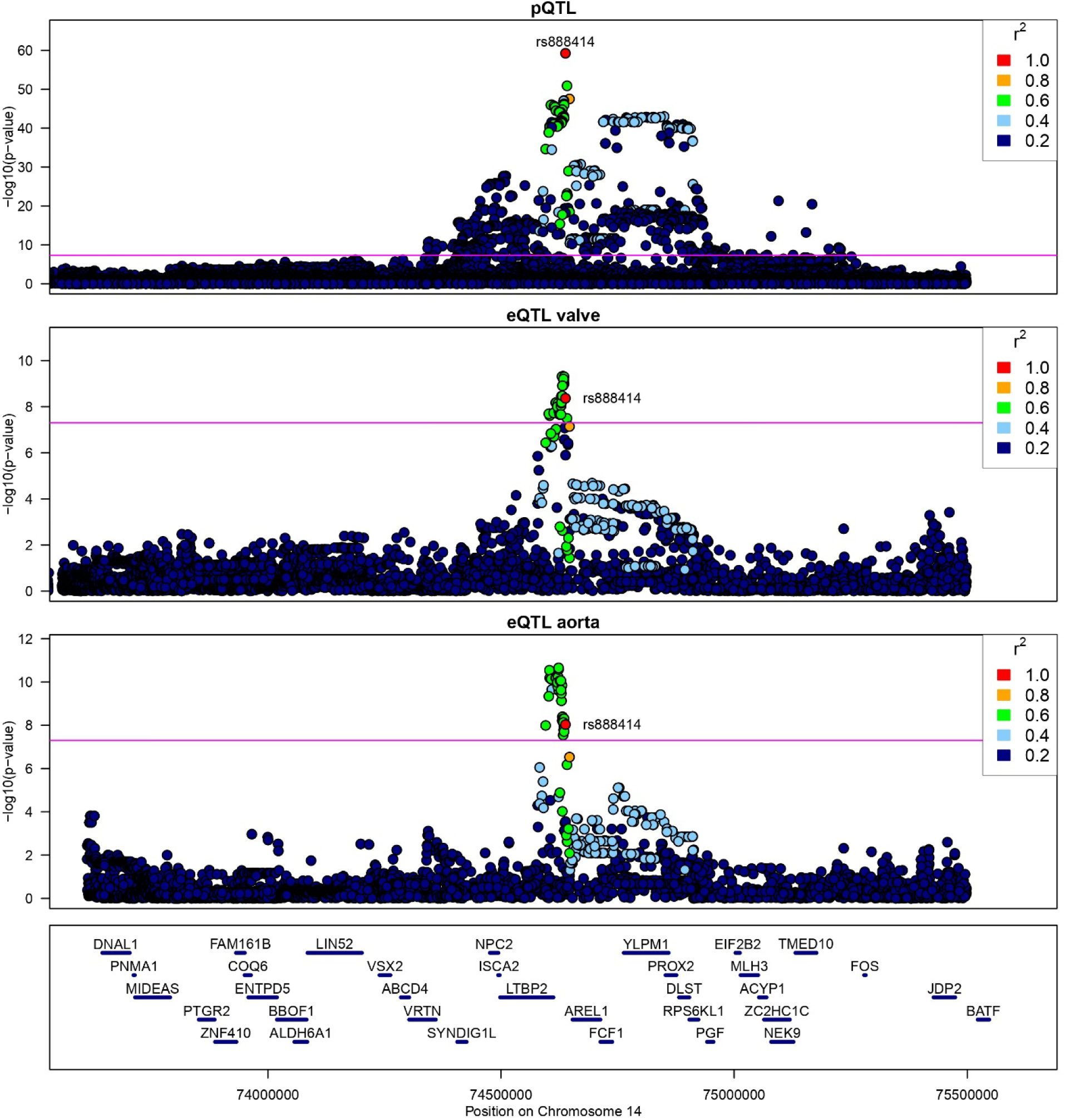
Association between genetic variants, plasma protein level and gene expression at the *LTBP2* locus. Regional plot representing the genetic associations across chromosome 14 (73.5-75.5 Mb) for circulating LTBP2 protein levels (pQTL, UKB-PPP), *LTBP2* expression in aortic valve tissues (eQTL, IUCPQ-ULaval cohort), and *LTBP2* expression in aorta (eQTL, GTEx v8). Points represent individual variants colored by linkage disequilibrium (r²) with the lead pQTL SNP (rs888414). Horizontal lines indicate genome-wide significance thresholds (p < 5E10-8).

### Protein Druggability

We verified druggability for the proteins associated with incident AS in the fully adjusted model with causal roles from pQTL or eQTL IVW MR analyses and no evidence of horizontal pleiotropy (REN, REG1A, CHI3L1, PCSK9, C7, NFASC, PRSS8, COL4A1, LTBP2, B2M).

Using DGIdb, six out of the ten prioritized genes showed high interaction scores, each exceeding the threshold of 0.1, suggesting potential for therapeutic intervention. Detailed interaction information, including drug type and approval status, is provided in Supplementary Table 11. We identified approved pharmacological agents targeting five of the prioritized protein biomarkers: B2M, COL4A1, PCSK9, REG1A, and REN.

## Discussion

In UK Biobank, a prospective cohort of adults from the United Kingdom with predominantly European ancestry, we identified circulating proteins significantly associated with AS risk, including sex-specific associations. Using aortic valve transcriptomic and MR, we prioritized potentially causal and specific candidates.

In the fully adjusted model, 55 proteins were associated with incident AS. Pathway enrichment analyses implicated inflammation, particularly TNF signaling, NF-κB activation, and apoptotic signaling. These findings align with prior evidence suggesting that chronic inflammation drives valvular calcification and sclerosis^24^. Our results confirm and extend a previous study using an earlier UKB-PPP release (1459 proteins, 44,313 individuals and 326 cases). Out of the 21 proteins identified in this study, we observed consistent associations for 13 proteins including GDF15, NT-proBNP, HAVCR1, TNFRSF9, IGFBP4, IL2RA, LTBP2, TNFRSF10A, PLAUR, IGFBP7, SPINK1, CD274, EDA2R^25^. GDF15 showed the strongest association (HR = 1.93 per SD increase). As a stress-responsive cytokine, GDF15 is elevated in many cardiac disorders and may reflect underlying valvular pathological processes driven by chronic inflammation and oxidative stress^26–28^. Another recent large-scale proteomic study in the Atherosclerosis Risk in Communities (ARIC) cohort (n=11,430) identified 52 plasma proteins associated with aortic stenosis progression, measured using the SomaScan platform. Their findings highlighted three key biomarkers showing consistent associations with aortic valve hemodynamics, calcification progression, and incident aortic stenosis events: C1QTNF1, GDF15, and MMP12^29^. Our analysis for incident AS confirmed the association for GDF15 and MMP12. Another important proteomic analysis for AS in the Cardiovascular Health Study and Age, Gene/Environment Susceptibility-Reykjavik Study, which also employed the SomaScan platform and adjudicated events, reported a distinct set of associated proteins^30^. The limited overlap with our findings could reflect methodological differences, including our focus on incident AS in a younger cohort (UK Biobank), and the use of a different proteomic platform (Olink).

We also report distinct sex-specific associations between circulating proteins and the incidence of AS, some of them showing significant interaction by sex. Five proteins had a stronger association in women (CD38, CD80, IGFBPL1, NFASC, and SERPINA9) whereas one had a stronger association in men (REG1A). CD38 was previously shown to promote cardiac hypertrophy and fibrosis via calcific calcium-NFAT signaling^31^. Immunohistochemical analysis of atherosclerotic plaques from human arteries, including the descending aorta, revealed CD80 expression in macrophages and a small subset of T cells, predominantly localized to the fibrous cap, thereby linking CD80 expression to the inflammatory processes underlying plaque development^32^. Plasma REG1A has been shown to be elevated in acute myocardial infarction, indicating a role in cardiovascular stress response^33^. The other identified proteins have not been previously linked to cardiovascular diseases. These sex-specific differences may reflect underlying biological mechanisms. Among women, the top enriched pathways included regulation of systemic arterial blood pressure, cellular response to oxidative stress, and diabetic cardiomyopathy. These pathways suggest that hemodynamic and metabolic stressors may play a more prominent role in AS development in females. In contrast, men demonstrated enrichment in pathways related to lipoprotein biosynthetic process, NABA ECM affiliated proteins, and actin filament-based movement. These findings are in line with prior evidence linking lipid metabolism and extracellular matrix remodeling to valve calcification, particularly in male individuals^9,34,35^. Taken together, our findings not only validate previous protein associations but also provide novel insights into sex-specific disease mechanisms, emphasizing the importance of personalized approaches to biomarker development and risk stratification in AS.

Complementing these observational findings, our Mendelian randomization analysis identified 16 proteins with putative robust causal effects, of which 4 were associated with incident AS in the fully adjusted model. PCSK9 showed the strongest association, in accordance with earlier work by Perrot et al. who demonstrated that carriers of the PCSK9 loss-of-function R46L variant had a significantly lower prevalence of AS, and that PCSK9 inhibition reduced calcium accumulation in human valvular interstitial cells^36^. These findings further align with the more recent MR analysis by Yang et al., which also reported a robust causal effect of plasma PCSK9 on AS in independent cohorts^37^. A significant causal effect was also observed for CHI3L1, a glycoprotein highly secreted by macrophages in inflammatory conditions^38^. Notably, it was previously found to be upregulated in human calcified aortic valves and associated with increased mortality in individuals with aortic stenosis^39,40^.

To prioritize proteins with a direct role in the target tissue, we identified those with high gene expression in the aortic valve. Leveraging tissue eQTLs using MR, we showed that genetically elevated aortic valve expression of *COL4A1*, *LTBP2*, and *B2M* increases AS risk. Colocalization analyses supported shared genetic regulation between *LTBP2* expression in valve and circulating protein levels. These findings suggest that LTBP2, a regulator of TGF-β signaling and fibrosis, may contribute directly to valvular pathology. As the largest member of the LTBP family, LTBP2 possesses unique N-terminal linker regions and shares structural homology with fibrillins, which are critical for extracellular matrix organization. Through an integrated multi-omics analysis of human right ventricular tissues and plasma, LTBP2 was identified as a blood biomarker upregulated in failing heart tissue of individuals with pulmonary artery hypertension, correlating with cardiac fibrosis^41^. In a different study, expression of LTBP2 was primarily localized within fibrotic regions of the myocardium during heart failure, suggesting its potential as a marker of cardiac fibrosis^42^. *COL4A1* encodes a portion of type IV collagen, a major component of the tissue basement membrane. Its expression in vascular smooth muscle cells is induced by TGFβ^43^. Genetic studies implicate the *COL4A1* locus in cardiovascular diseases. A variant in *COL4A1* was associated with increased arterial stiffness^44^. Variants in *COL4A2* affect *COL4A1*/*COL4A2* expression, leading to smooth muscle cell apoptosis, thinner fibrous caps, and higher myocardial infarction risk^45^. B2M (β-2-microglobulin), a component of major histocompatibility complex (MHC) class I, is a polypeptide present on the surface of nucleated human cells. B2M is associated with antigen presentation, inflammation, and the complement cascade, with elevated levels linked to cardiovascular events and mortality^46^. Our observation that genetically predicted B2M expression increases AS risk aligns with its association with systemic inflammation, though the lack of colocalization between pQTLs and aortic valve eQTLs suggests the effects may be mediated through broader inflammatory pathways rather than valve-specific mechanisms.

Our study identified several druggable targets for AS. PCSK9 showed the strongest causal association in blood and is already targeted by approved monoclonal antibodies (alirocumab, evolocumab) and a small interfering RNA (inclisiran), suggesting immediate repurposing potential. This hypothesis is currently being tested in an ongoing randomized controlled trial evaluating the effect of PCSK9 inhibition on the progression of aortic stenosis (NCT04968509). Another randomized controlled trial with the same objective is planned (NCT06996223). REN is directly targeted by renin–angiotensin inhibitors and clinical observations report that renin–angiotensin system blockade is associated with slower progression of aortic stenosis, reduction in left ventricular mass, and significant survival benefits in patients undergoing aortic valve replacement^47,48^. A recent pilot case-control study demonstrated that angiotensin receptor blockers (ARBs) significantly slow aortic valve calcification progression in patients with mild to moderate aortic stenosis, with the strongest benefit observed in women^49^. Moreover, a randomized controlled trial is ongoing (NCT04913870). LTBP2 currently lacks direct inhibitors but could be approached indirectly through TGF-β pathway modulation. These findings highlight both repurposing opportunities and avenues for novel therapeutic development.

### Limitation

Several limitations should be acknowledged. First, our analysis focused mainly on individuals of European ancestry (93.2%), limiting generalizability to other populations. Second, due to the unavailability of sex-stratified GWAS summary statistics, we were unable to perform MR analyses separately in women and men. Instead, MR analyses were conducted in a combined sample, which may obscure sex-specific effects at the genetic level. Third, while our MR approach strengthens causal inference, functional validation and mechanistic studies are required to confirm these findings in vitro and in vivo. Future directions should include expanding analyses to multi-ancestry cohorts and investigating the utility of the identified proteins for early detection or risk stratification.

## Conclusion

By integrating observational analyses with causal inference methods to identify both shared and sex-specific disease blood biomarkers, we identified aortic valve-specific proteins strongly associated with the incidence of AS. Together, our results highlight the utility of proteogenomic approaches for understanding disease pathogenesis, biomarker discovery and identification of therapeutic targets.

## Data Availability

The RNA-seq data have been deposited at dbGaP as dbGaP: phs003541.v1.p1. They are available upon request if access is granted. Access can be obtained for research related to cardiovascular diseases by not-for-profit organizations providing a local institutional review board approval and a letter of collaboration with the study investigators. Requests can be made to the corresponding author.

https://gtexportal.org/home/datasets

