## Supplementary Figures 1-3 for "Connecting proteomics and genomics to identify causal biomarkers specific for aortic valve stenosis"

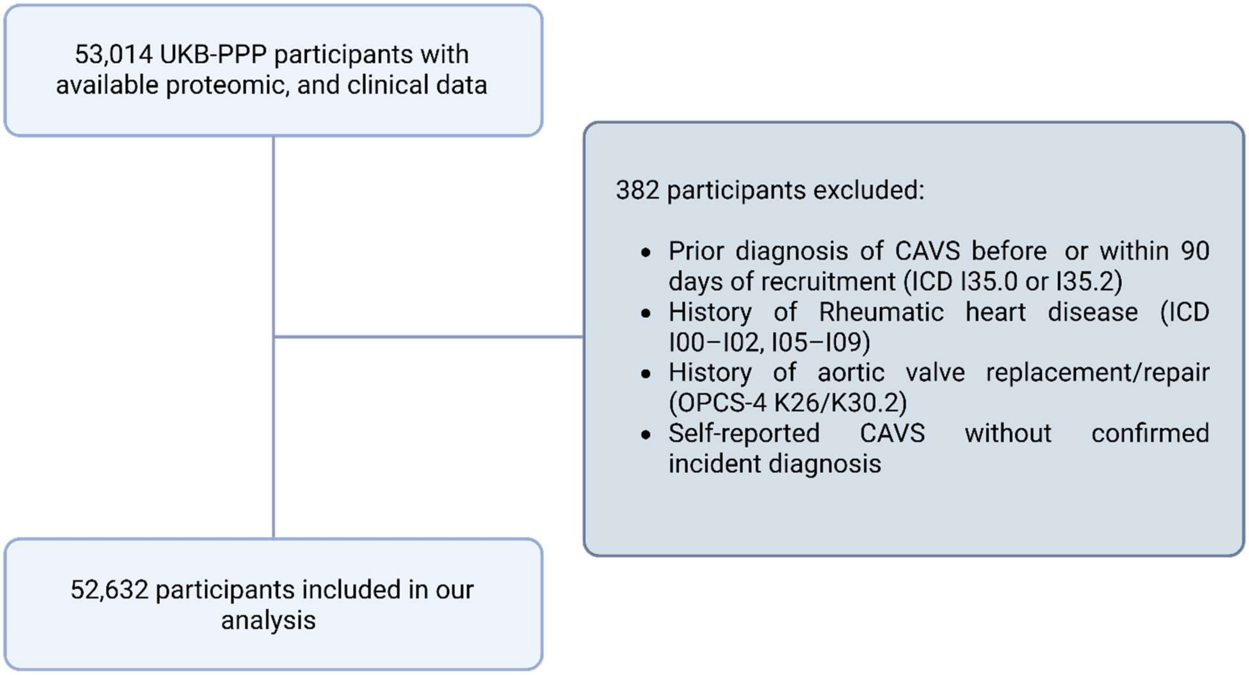


**Supplementary Figure 1.** Overview of participants selection and eligibility criteria.

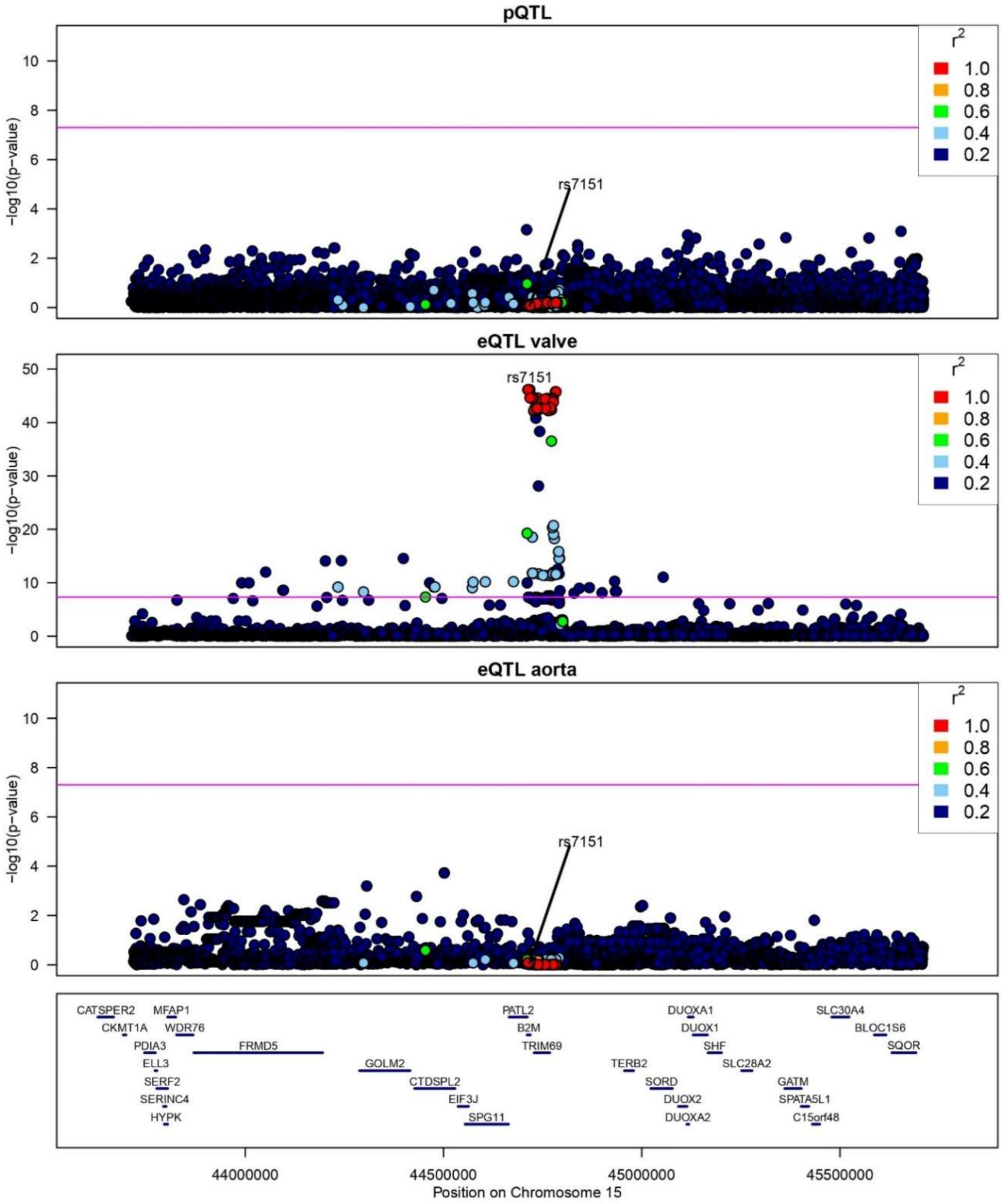


**Supplementary Figure 2.** Regional plot for B2M. Genetic associations across chromosome 15 (43.7-45.7 Mb) for circulating B2M protein levels (pQTL, UKB-PPP), *B2M* expression in aortic valve tissue (eQTL, IUCPQ-ULaval cohort), and aorta expression (eQTL, GTEx v8). Points represent individual variants colored by linkage disequilibrium ($r^2$) with the lead SNP (rs7151). Horizontal lines indicate genome-wide significance thresholds ($p < 5E-8$).

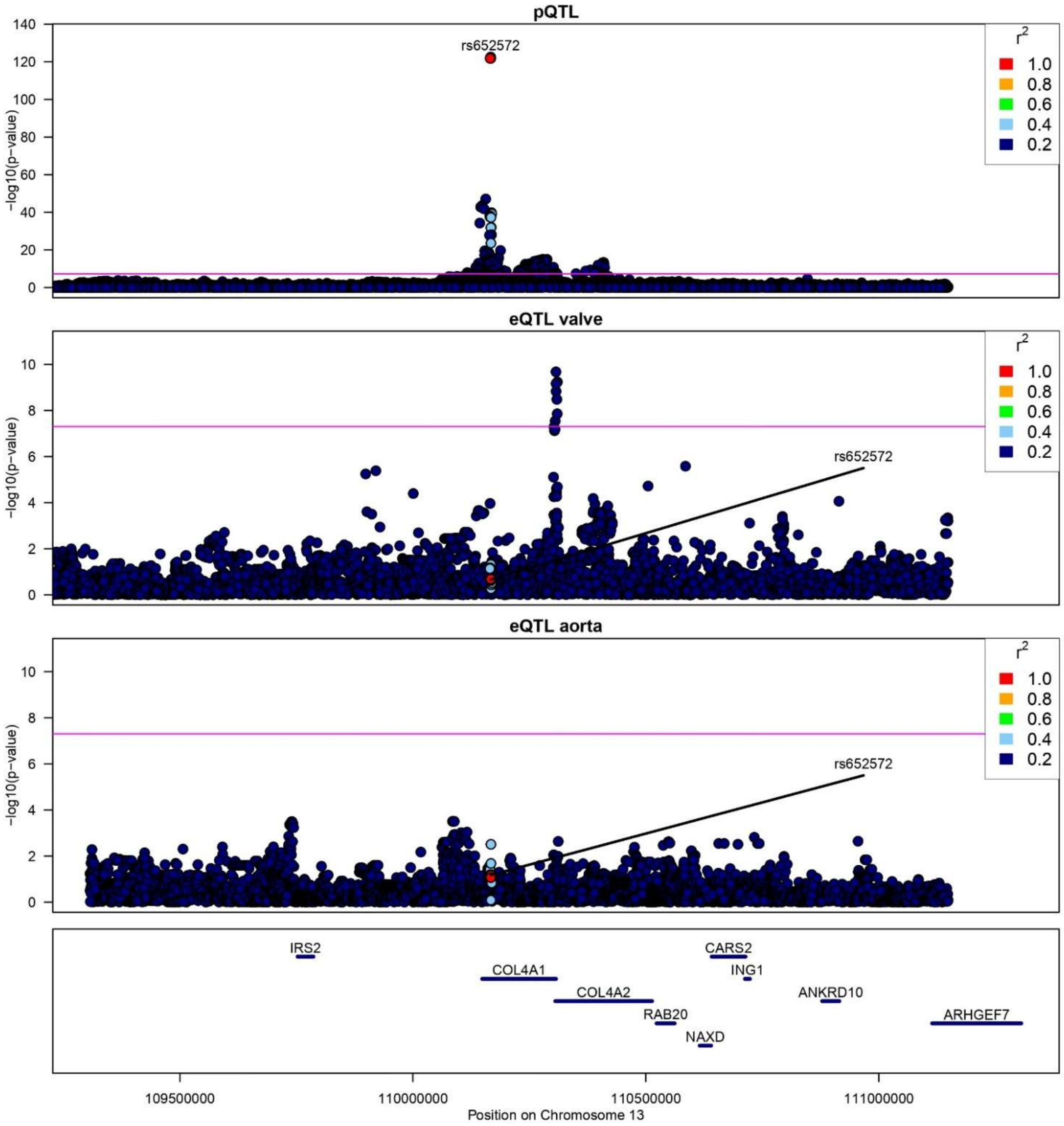


**Supplementary Figure 3.** Regional plot for COL4A1. Genetic associations across chromosome 13 (109.1-111.1 Mb) for circulating COL4A1 protein levels (pQTL, UKB-PPP), *COL4A1* expression in aortic valve tissue (eQTL, IUCPQ-ULaval cohort), and aorta expression (eQTL, GTEx v8). Points represent individual variants colored by linkage disequilibrium ($r^2$) with the lead SNP (rs652572). Horizontal lines indicate genome-wide significance thresholds ($p < 5E-8$).
